# Age-specific effects of childhood Body Mass Index on multiple sclerosis risk

**DOI:** 10.1101/2021.09.12.21263445

**Authors:** Luke Hone, Benjamin M Jacobs, Charles Marshall, Gavin Giovannoni, Alastair Noyce, Ruth Dobson

## Abstract

**Objective:** Higher Body Mass Index (BMI) during early life is thought to be a causal risk factor for Multiple Sclerosis (MS). We used longitudinal mendelian randomisation (MR) to determine whether there is a critical window during which BMI influences MS risk.

**Methods:** Summary statistics for childhood BMI and for MS susceptibility were obtained from recent large GWAS. We generated exposure instruments for BMI during four non-overlapping epochs (< 3 months, 3 months-1.5 years, 2-5 years, and 7-8 years) and performed MR using the inverse-variance weighted method with standard sensitivity analyses. Multivariable MR was used to account for effects mediated via later-life BMI.

**Results:** For all time epochs other than birth, genetically determined higher BMI was associated with an increased liability to MS: Birth (OR 0.81, 95% CI 0.50-1.31, N_SNPs_=7, p=0.39), Infancy (OR 1.18, 95%CI 1.04-1.33, N_SNPs_=18, p=0.01), Early childhood (OR 1.31, 95%CI 1.03-1.66, N_SNPs_=4, p=0.03), Later childhood (OR 1.34, 95% CI 1.08-1.66, N_SNPs_=4, p=0.01). Multivariable MR suggested that these effects may be mediated by effects on adult BMI.

**Conclusion:** We provide evidence using MR that genetically-determined higher BMI during early life is associated with increased MS risk. This effect may be driven by shared genetic architecture with later-life BMI.

## Introduction

Multiple Sclerosis (MS) is an immune-mediated demyelinating disease of the Central Nervous System (CNS), with both genetic and environmental risk factors^1^. Environmental factors including obesity, low serum vitamin D, EBV infection, smoking, air pollution and solvent exposure have all been associated with increased MS risk, although the mechanisms through which they act remain uncertain^1^. Traditional epidemiological study designs used to study environmental risk factors - such as case-control and cohort studies - are liable to unmeasured confounding and reverse causation, yielding associations which are not truly causal^2^.

Mendelian Randomisation (MR) is a type of instrumental variable (IV) analysis that uses genetic variants as proxies to examine whether observed associations between an exposure and an outcome are likely to be causal^2,3^. Childhood obesity has been identified as a potential causal MS risk factor in both epidemiological^4,5^ and MR studies^5–7^. A consistent finding is that birth weight is not associated with MS risk, whereas higher BMI during later childhood and adolescence appears to be a risk factor^4,7–9^. However, BMI is a dynamic trait; it is unknown if there is a crucial point during development when BMI influences subsequent MS risk. Understanding the precise nature and timing of this association may shed light on the biological pathways leading to the clinical development of MS and may inform targeted obesity prevention programmes in early life. Defining this critical window of effect is challenging using MR to instrument BMI at a single time point. Although genetic instruments for BMI throughout the life course can be obtained from different datasets, using GWAS from a single, longitudinal cohort minimises the risk of subtle population stratification influencing the results.

Longitudinal MR analysis of the effect of BMI during development is possible due to the recent publication of longitudinal BMI GWAS data from the Norwegian Mother, Father, and Child Cohort Study (MoBa) cohort^11^. We set out to explore the causal effect of BMI changes during early life on MS risk and examine the dynamics of this effect over time.

## Methods

### Genetic datasets

#### Exposures dataset

The MoBa cohort is an open-ended cohort study that recruited pregnant women in Norway from 1999 to 2008, with anthropometric measurements of the children by trained nurses across childhood. This cohort was used to perform GWAS of BMI at various ages using data from between 11,095 and 28,681 children across 12 time points from birth to 8 years (birth, 6 weeks, 3 months, 6 months, 8 months, 1 year, 1.5 years, 2 years, 3 years, 5 years, 7 years, and 8 years). 46 distinct genetic loci were associated with higher childhood BMI at various ages^11^. The outcome of this GWAS was standardized BMI, and therefore the units are not directly comparable with published estimates from MR studies using other BMI GWAS for genetic instruments.

We used variants reaching a genome-wide p-value threshold of <5×10^−8^ at each time point after conditional and joint analysis as instrumental variables. In the primary analysis, time points were amalgamated into four distinct epochs: Birth to 6 weeks, 3 months to 1.5 years, 2 to 5 years, and 7 to 8 years, reflecting the developmental stages of birth, infant, toddler and child. If a variant was associated at p<5×10^−8^ with >1 time point within an epoch, the association statistics were taken from the time point with the strongest association (i.e. the smallest p value). A secondary analysis then used data at each individual time point. These epochs were defined according to the shared genetics of BMI at each time point, and loosely correspond to the four phases of BMI genetics suggested by the authors^11^. Data are available from the MoBa authors on request.

### Outcome dataset

The International Multiple Sclerosis Genetics Consortium (IMSGC) 2019 discovery phase GWAS (47,429 MS cases, 68,374 controls) was used as the outcome dataset^11^. We used summary statistics from the discovery stage meta-analysis (14,802 persons with MS, 26,703 controls). This GWAS discovered 233 independent genome-wide significant signals associated with MS, of which 32 variants lay within the Major Histocompatibility Complex (MHC). Data are available on request from https://imsgc.net/.

### Other datasets

GWAS datasets for adult BMI and birthweight were obtained from the GIANT and EGG consortia meta-analyses (respectively) including UK Biobank participants^12,13^. Data on birth weight has been contributed by the EGG Consortium using the UK Biobank Resource and has been downloaded from www.egg-consortium.org. Data on adult BMI can be downloaded from https://portals.broadinstitute.org/collaboration/giant/index.php/GIANT_consortium_data_files.

### Mendelian randomisation & statistical analyses

Mendelian Randomisation (MR) was performed using the TwoSampleMR (version 0.5.5) package in R (version 3.6.1)^14^.

Genetic instruments were generated by the following steps:

1. Exposure (BMI) Single Nucleotide Polymorphisms (SNPs) associated with childhood BMI at p<5×10^−8^ were retained.
2. SNPs were clumped to ensure independence using the PLINK clumping method and a European 1000 Genomes reference dataset. SNPs in linkage disequilibrium (LD) (R^2^> 0.001) or within a window of 10,000 kb were pruned. The SNP with the lowest p-value in the window or in LD was retained for analysis.
3. MHC SNPs (chr6:25,000,000 - chr6:35,000,000 in hg19) were excluded.
4. Exposure SNPs were extracted from the outcome dataset (IMSGC MS GWAS).
5. Exposure and outcome SNPs were harmonised so that their effects corresponded to the same allele and palindromic SNPs with a minor allele frequency (MAF) > 0.42 were discarded.

MR analysis was performed using a random-effects, inverse-variance-weighted method (IVW). Secondary analyses included a test for heterogeneity, tests for horizontal pleiotropy through calculation of an MR-Egger intercept and MR-PRESSO^15^, and a leave-one-out analysis. In secondary analyses we excluded SNPs explaining more variance in the outcome than exposure (Steiger filtering) as these variants are likely to violate the exclusion-restriction assumption of MR^16^. Results are presented as Odds Ratios (OR) with 95% Confidence Intervals (CIs) and associated p-values. Results are visualised as scatter, forest, funnel and leave-one-out plots. F-statistics for instrument strength were calculated for each SNP as β^2^/SE^2^, where β is the beta coefficient estimating the per-allele effect on BMI, and SE is the standard error of the beta coefficient. Mean F statistics for each epoch were calculated and are displayed in supplementary table 1. All mean F statistics were above 10, suggestive of strong instruments.

To determine the extent to which the observed effect of genetically-estimated higher early life BMI on MS risk was mediated by adult BMI, we performed multivariable MR, conditioning on the effect of each instrument on adult BMI. Adult BMI GWAS summary statistics were obtained from the GIANT consortium GWAS meta-analysis^12^. SNPs in the childhood BMI instrument were first harmonised with the adult BMI GWAS. After harmonisation, only one variant remained for the 2 years – 5 years epoch, precluding multivariable analysis. For other epochs, we performed multivariable MR using the residual-based method as described in Burgess *et al* (2015)^17^. In brief, this is a two-step approach which first regresses SNP associations with the outcome (MS) on SNP associations with the secondary risk factor (adult BMI). The residuals from this first step represent the variation in the outcome not explained by the secondary risk factor. These residuals are then regressed on the main exposure of interest (childhood BMI), giving an estimate of the effect of this exposure on the outcome independent of the secondary risk factor. As a sensitivity analysis, we also performed multivariable MR by jointly modelling the effects of SNPs on both risk factors in a weighted regression (implemented in *mv_multiple* in TwoSampleMR). As a further sensitivity analysis, we repeated the univariable MR for each time epoch, restricting the instruments to SNPs for which a) associations with adult BMI were reported in the GIANT GWAS and b) these associations were weaker than an arbitrary P value cutoff (P>0.05). The intuition behind this approach is that, given the power of the GIANT GWAS, removing SNPs with even weak evidence of association with adult BMI (e.g. P<0.05) should restrict the genetic instrument to those SNPs with an effect on childhood BMI, but not later-life BMI.

To provide further confirmation of the birthweight MR result, we repeated the analysis using an independent and larger GWAS of birthweight including UKB participants^13^.

### Data Availability

We thank MoBa, Prof Stefan Johansson and Dr Marc Vaudel for providing summary statistics for childhood BMI. We thank the IMSGC for providing MS GWAS data. We thank MR-Base for making the TwoSampleMR package available publicly. Download links for datasets used in this study are provided above. All code used in this study is available at https://github.com/benjacobs123456/MR_MOBA_MS.

### Consent and Approval

This study was performed using publicly available data sources. There were no experiments performed on human subjects and no direct patient contact, therefore written consent was not required.

## Results

### Age-epoch analysis

We collapsed BMI during childhood into four windows: birth (birth - 6 weeks inclusive), infancy (3 months - 1.5 years inclusive), early childhood (2 years - 5 years inclusive), and later childhood (7 - 8 years). For all time epochs, other than birth, genetically-estimated higher BMI was associated with an increased liability to MS (tables 1 & 2, figure 1, supplementary figures 1&2): birth (OR 0.81, 95% CI 0.50 - 1.31, N_SNPs_=7, p=0.39), infancy (OR 1.18, 95% CI 1.04 - 1.33, N_SNPs_=18, p = 0.01), early childhood (OR 1.31, 95% CI 1.03 - 1.66, N_SNPs_=4, p=0.03), and later childhood (OR 1.34, 95% CI 1.08 - 1.66, N_SNPs_=4, p=0.01).

**Table 1:** Single Nucleotide Polymorphisms (SNPs) associated with BMI at various time epochs during early life. ‘Epoch’ refers to the time window in question. For each SNP, the nearest gene, and the beta, standard error (SE) and associated P value are given for the association with standardized BMI (at each time point) and with MS (i.e. the beta is the log odds ratio per effect allele copy). EAF – effect allele frequency. BMI – body mass index. SNP – single nucleotide polymorphism. SE – standard error. MS – multiple sclerosis.

| Epoch | SNP | Nearest Gene | Effect allele | Non-effect allele | EAF in MOBA | Exposure (BMI) |  |  | Outcome (MS) |  |  |
| --- | --- | --- | --- | --- | --- | --- | --- | --- | --- | --- | --- |
|  |  |  |  |  |  | Beta | SE | P value | Beta | SE | P value |
| Birth - 6 weeks | rs11187129 | *HHEX* | T | C | 0.54108 | -0.0471726 | 0.008338 | 2.10E-08 | 0.094401 | 0.01739 | 5.68E-08 |
| Birth - 6 weeks | rs11708067 | *ADCY5* | A | G | 0.765083 | -0.0789424 | 0.009755 | 5.20E-16 | -1.00E-04 | 0.029553 | 0.9973 |
| Birth - 6 weeks | rs1482853 | *CCNLI* | C | A | 0.598464 | 0.0988178 | 0.008435 | 5.90E-32 | -0.01511 | 0.016958 | 0.3728 |
| Birth - 6 weeks | rs2298615 | *EHBP1LI* | C | T | 0.76651 | -0.0712846 | 0.012196 | 5.40E-09 | 0.045929 | 0.02142 | 0.03202 |
| Birth - 6 weeks | rs28642213 | *GPSMI* | A | G | 0.267403 | 0.0623516 | 0.009521 | 4.70E-11 | 0.02245 | 0.028404 | 0.4293 |
| Birth - 6 weeks | rs7310615 | *SH2B3* | C | G | 0.45479 | -0.0498278 | 0.008508 | 6.50E-09 | 0.067752 | 0.016702 | 4.98E-05 |
| Birth - 6 weeks | rs739669 | *DLG4* | G | A | 0.376996 | -0.0715704 | 0.008601 | 4.70E-17 | -0.02358 | 0.01775 | 0.1841 |
| Birth - 6 weeks | rs7772579 | *ESR1* | A | C | 0.699604 | 0.0651087 | 0.009067 | 5.90E-13 | 0.002297 | 0.018227 | 0.8997 |
| 3 months - 1.5 years | rs1032296 | *HHIP* | T | C | 0.376232 | 0.0524932 | 0.009097 | 1.10E-08 | 0.025831 | 0.017131 | 0.1316 |
| 3 months - 1.5 years | rs10493544 | *TNNI3K* | T | C | 0.430744 | 0.0538075 | 0.009404 | 7.10E-09 | -0.00638 | 0.01639 | 0.6971 |
| 3 months - 1.5 years | rs11676272 | *ADCY3* | A | G | 0.51411 | -0.0887022 | 0.009299 | 2.80E-22 | -0.0203 | 0.016619 | 0.2218 |
| 3 months - 1.5 years | rs13038017 | *EFCAB8* | T | C | 0.47431 | -0.0535926 | 0.009372 | 1.20E-08 | 0.001601 | 0.016545 | 0.9229 |
| 3 months - 1.5 years | rs17145750 | *MLXIPL* | C | T | 0.840536 | 0.0697872 | 0.011991 | 6.80E-09 | -0.02706 | 0.025971 | 0.2974 |
| 3 months - 1.5 years | rs1772945 | *OPRM1* | G | A | 0.441547 | -0.0562618 | 0.00938 | 3.20E-09 | -0.0031 | 0.016224 | 0.8487 |
| 3 months - 1.5 years | rs1820721 | *GLPR1* | C | A | 0.511425 | -0.060729 | 0.008743 | 7.20E-12 | 0.016333 | 0.016275 | 0.3156 |
| 3 months - 1.5 years | rs1985927 | *SCGB1A1* | T | C | 0.270127 | -0.0597242 | 0.010637 | 6.80E-09 | -0.0244 | 0.019192 | 0.2036 |
| 3 months - 1.5 years | rs209421 | *UBE3D* | G | T | 0.264271 | 0.0727381 | 0.009951 | 5.40E-13 | 0.006521 | 0.018044 | 0.7178 |
| 3 months - 1.5 years | rs2585058 | *SH3GL3* | G | A | 0.525095 | 0.0630845 | 0.009321 | 8.60E-12 | 0.007373 | 0.016718 | 0.6592 |
| 3 months - 1.5 years | rs2728641 | RP11-405A12.2 | C | T | 0.477989 | 0.0501517 | 0.008951 | 1.90E-08 | 0.0244 | 0.016829 | 0.1471 |
| 3 months - 1.5 years | rs2767486 | *LEPR* | A | G | 0.838584 | -0.14325 | 0.011859 | 6.40E-34 | -0.03816 | 0.021062 | 0.07 |
| 3 months - 1.5 years | rs2816985 | *NR5A2* | A | G | 0.551004 | -0.0591627 | 0.009057 | 5.40E-11 | -0.02762 | 0.016419 | 0.09259 |
| 3 months - 1.5 years | rs28457693 | *PTCH1* | A | G | 0.865473 | -0.0726875 | 0.01297 | 2.40E-08 | -0.03449 | 0.029121 | 0.2363 |
| 3 months - 1.5 years | rs287621 | *KLF14* | T | C | 0.260505 | 0.0636843 | 0.010075 | 3.70E-10 | -0.0141 | 0.018164 | 0.4376 |
| 3 months - 1.5 years | rs3741508 | *NCOR2* | T | G | 0.856761 | 0.0826905 | 0.013414 | 1.20E-09 | 0.009356 | 0.025142 | 0.7098 |
| 3 months - 1.5 years | rs6538845 | RP11-690J15.1 | T | C | 0.515215 | -0.0549436 | 0.008957 | 1.50E-09 | -0.02567 | 0.016303 | 0.1154 |
| 3 months - 1.5 years | rs6899303 | *PCSK1* | A | C | 0.368755 | -0.057154 | 0.009025 | 5.30E-11 | -0.00409 | 0.016591 | 0.8052 |
| 2 years - 5 years | rs11676272 | *ADCY3* | A | G | 0.515086 | -0.0750482 | 0.010748 | 2.70E-12 | -0.0203 | 0.016619 | 0.2218 |
| 2 years - 5 years | rs12672489 | *KLF14* | C | T | 0.745792 | 0.0726685 | 0.012849 | 1.10E-08 | 0.015775 | 0.018633 | 0.3972 |
| 2 years - 5 years | rs1830890 | *PLCE1* | A | G | 0.678614 | -0.0665976 | 0.011646 | 1.30E-08 | -0.00733 | 0.017525 | 0.6759 |
| 2 years - 5 years | rs2767486 | *LEPR* | A | G | 0.837712 | -0.0853349 | 0.014622 | 4.60E-09 | -0.03816 | 0.021062 | 0.07 |
| 7 years - 8 years | rs10493544 | *TNNI3K* | T | C | 0.43154 | 0.0651591 | 0.01198 | 4.80E-08 | -0.00638 | 0.01639 | 0.6971 |
| 7 years - 8 years | rs11676272 | *ADCY3* | A | G | 0.515866 | -0.078168 | 0.013323 | 2.90E-09 | -0.0203 | 0.016619 | 0.2218 |
| 7 years - 8 years | rs17817288 | *FTO* | A | G | 0.513158 | -0.0952937 | 0.013447 | 1.30E-12 | -0.03325 | 0.016187 | 0.03999 |
| 7 years - 8 years | rs545608 | *SEC16B* | G | C | 0.766209 | -0.0877785 | 0.015795 | 3.20E-08 | -0.01114 | 0.020384 | 0.5848 |
| 7 years - 8 years | rs7132908 | *FAIM2* | G | A | 0.598895 | -0.0813492 | 0.01358 | 3.30E-09 | -0.03999 | 0.016912 | 0.01805 |

**Table 2:** MR estimates from the primary analysis of BMI during each time epoch on MS. For each method, the number of included SNPs is included. The IVW (inverse variance weighted) method was used as the primary analytic method. MR – mendelian randomisation.

| Exposure epoch | MR method | Number of SNPs | Beta | SE | P value |
| --- | --- | --- | --- | --- | --- |
| Birth - 6 weeks | Inverse variance weighted | 7 | -0.20891 | 0.243413 | 0.390749 |
| 3 months - 1.5 years | Inverse variance weighted | 18 | 0.162189 | 0.062848 | 0.009861 |
| 2 years - 5 years | Inverse variance weighted | 4 | 0.267068 | 0.122666 | 0.029467 |
| 7 years - 8 years | Inverse variance weighted | 4 | 0.289007 | 0.109746 | 0.008453 |
| Birth - 6 weeks | MR Egger | 7 | 1.184225 | 0.981039 | 0.281374 |
| 3 months - 1.5 years | MR Egger | 18 | 0.259406 | 0.223306 | 0.262409 |
| 2 years - 5 years | MR Egger | 4 | 1.652619 | 1.447985 | 0.371971 |
| 7 years - 8 years | MR Egger | 4 | 1.340814 | 0.760078 | 0.219773 |
| Birth - 6 weeks | Simple mode | 7 | 0.091193 | 0.189569 | 0.647517 |
| 3 months - 1.5 years | Simple mode | 18 | 0.10474 | 0.14939 | 0.492715 |
| 2 years - 5 years | Simple mode | 4 | 0.219738 | 0.197409 | 0.346823 |
| 7 years - 8 years | Simple mode | 4 | 0.349886 | 0.181151 | 0.148952 |
| Birth - 6 weeks | Weighted median | 7 | -0.0515 | 0.145521 | 0.723423 |
| 3 months - 1.5 years | Weighted median | 18 | 0.179452 | 0.086761 | 0.038606 |
| 2 years - 5 years | Weighted median | 4 | 0.248544 | 0.140316 | 0.076509 |
| 7 years - 8 years | Weighted median | 4 | 0.320545 | 0.127328 | 0.01182 |
| Birth - 6 weeks | Weighted mode | 7 | -0.03037 | 0.149958 | 0.846181 |
| 3 months - 1.5 years | Weighted mode | 18 | 0.196361 | 0.116829 | 0.11109 |
| 2 years - 5 years | Weighted mode | 4 | 0.231029 | 0.189487 | 0.309874 |
| 7 years - 8 years | Weighted mode | 4 | 0.347357 | 0.165152 | 0.126174 |

**Figure 1:**
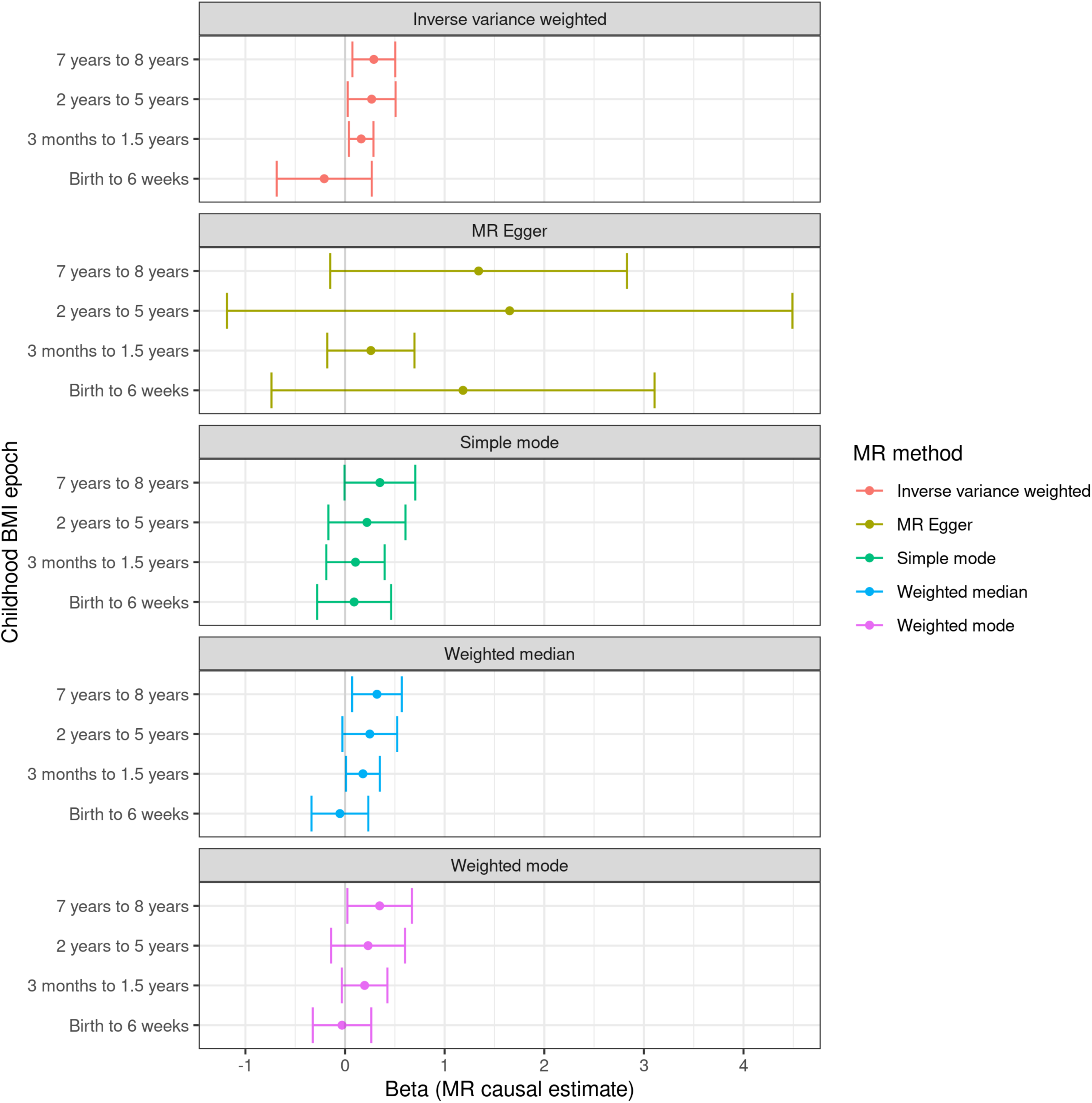
Forest plots showing the MR effect estimates during each epoch for the effect of BMI on MS susceptibility. Points represent beta estimates reflecting the predicted log odds ratio for MS risk per 1 unit increase in genetically-determined standardised BMI at each time point. Error bars show 95% confidence intervals. The separate panels display the primary MR analytic method – the inverse-variance weighted (IVW) method – followed by secondary sensitivity analyses.

The MR-Egger intercepts did not suggest that unbalanced horizontal pleiotropy was biasing the IVW estimate at any of these epochs (supplementary table 1). Cochran’s test of heterogeneity suggested heterogeneity in the IVW estimators during the birth epoch, but not during any other time window (supplementary table 1). Using the MR-PRESSO method, we found evidence of global pleiotropy for the birth epoch (p<0.001) but no other epochs (infancy p=0.68, early childhood p=0.84, later childhood p=0.43). For the birth epoch, a single outlier (rs11187129) was found to distort the overall MR estimate (P_Outlier_< 0.05), and removal of this outlier led to a point estimate closer to the null (beta_corrected_ = -0.06, p=0.69).

To determine whether these results could be driven by SNPs acting in the opposite causal direction (i.e. influencing childhood BMI via susceptibility to MS), we excluded any SNPs explaining more variance in the outcome than the exposure (Steiger filtering)^16^. No SNPs were filtered out using this approach, and thus the causal estimates were unchanged. Leave-one-out sensitivity analysis, in which SNPs are removed in turn from the genetic instrument to ensure no one SNP is driving the result, did not lead to an appreciable change in the MR estimates, although there was some loss of precision (supplementary table 2).

### The effect of early life BMI on MS risk is contingent on persistence of elevated BMI into later life

Next, we explored whether the observed MR effects of childhood BMI during infancy, early and late childhood on MS risk could be driven by the effects of these genetic variants on BMI in adulthood. To address this question, we performed multivariable MR, conditioning on the effect of each childhood BMI instrument on adult BMI, allowing us to estimate the ‘direct’ effect of elevated BMI at each time point independent of the effect mediated via adult BMI. Associations between SNPs and adult BMI were obtained from a large GWAS of ∼700,000 adults^12^. After conditioning on adult BMI, the causal estimates for each epoch diminished in magnitude, and did not show strong evidence for association with MS risk. This finding suggests that the observed effect at stage 1 may be due to persistence of higher BMI into later life (supplementary figure 3, supplementary tables 3&4). There is some loss of precision and power in these estimates compared to the univariable MR, largely because some variants were not included in the GIANT GWAS. For the early childhood epoch, only one SNP was retained and so the Wald Ratio method was the only method available to carry out the MR analysis. This attenuation of effect size was most marked for the later childhood epoch (univariable MR estimate OR 1.34, 95% CI 1.08 – 1.66, p=0.01; multivariable MR estimate OR 0.98, 95% CI 0.74 – 1.30, p=0.9).

To confirm these results, we repeated the univariable MR analysis using SNPs with no evidence of association with adult BMI. Specifically, we included SNPs for which a) summary statistics for association with adult BMI were available and b) were not associated with adult BMI at an arbitrary significance threshold (P<0.05). This lenient threshold was chosen to ensure that variants with even a weak effect on adult BMI were excluded. After excluding SNPs associated with adult BMI at P<0.05, no SNPs remained for the early or late childhood epochs, underlining the high correlation between BMI in these epochs and adult BMI. For the infancy epoch, although there was a loss of precision, the IVW estimate suggested a possible residual effect of elevated BMI at this time point (OR 1.28, 95% CI 0.97 – 1.68, p=0.08, N_SNP_=4), with no significant unbalanced horizontal pleiotropy biasing this estimate (Egger intercept 0.05, p=0.60). Each of these four SNPs - rs209421 (*UBE3D*), rs2816985 (*NR5A2*), rs2268657 (*GLP1R*) and rs1772945 (*OPRM1*) - influences BMI during infancy, with minimal effect on birth weight or subsequent BMI, and no direct association with MS risk (association P values all >0.05).

### Individual time point analysis

Secondary analysis at each time point demonstrated results consistent with the epoch-based analysis, albeit with less precision at each individual time point due to the smaller number of variants used for the genetic instruments (supplementary figure 4, supplementary table 5). A Mann-Kendall trend test of the 12 individual time points suggested evidence of a trend in the MR effect estimates (IVW or Wald Ratio where only one SNP was available for analysis; tau = 0.636, 2-sided p-value =0.005)

Although there was no evidence of unbalanced horizontal pleiotropy at any of these time points where there were sufficient SNPs in the instrument to test, there was again evidence of notable heterogeneity at the birth BMI time point (supplementary table 6).

To replicate our negative finding that birth weight does not causally impact MS risk using an external dataset, we repeated the analysis using summary statistics from a larger GWAS of birthweight^13^. This yielded a convincing null (OR 1.20, 95% CI 0.92 - 1.55, N_SNP_=136, p=0.17) with no evidence to suggest unbalanced horizontal pleiotropy (Egger-intercept = -0.01, p=0.18).

## Discussion

Using two-sample mendelian randomisation and longitudinal data from a Norweigan early life cohort, we replicate the finding that early life BMI appears to be a causal risk factor for Multiple Sclerosis. We find no evidence for an effect of genetically-estimated birth weight on subsequent MS risk. We report an increase in magnitude of the effect size of genetically-estimated higher BMI across the life course from early to late childhood. Using multivariable MR to control for the effects of these genetic variants mediated via effects on later-life BMI, we find that this gradient is likely to represent, in the main, an increasingly strong genetic correlation between BMI at each age epoch and late-adolescent/adult BMI. By removing SNPs associated with adult BMI, we find that some of the effect of elevated BMI during infancy may be independent of later-life BMI, however these results were imprecise and not supported in a multivariable analysis. It therefore seems likely that most of the causal effect of elevated early life BMI on MS risk is likely to depend on persistence of elevated BMI through to adolescence/early adulthood.

Here we attempt to dissect the time-varying effects of elevated BMI in childhood on MS risk using two-sample MR. Using longitudinal GWAS from the same cohort provides a unique opportunity to understand time-varying exposures, and is likely to minimise population stratification compared to using multiple different exposure GWAS from different time points in different cohorts. We use a variety of methods to quantify and account for pleiotropy, and demonstrate using the MR-PRESSO method that pleiotropy is unlikely to influence the positive results we observe at the 3 months - 1.5 years, 2 years - 5 years, and 7 years - 8 years epochs. We demonstrate the possible value of this approach for unpicking the role of time-varying factors on disease risk, demonstrating a clear trend from a null effect of birthweight to a clear effect of BMI in late childhood, consistent with previous epidemiological findings in MS^7,9^.

Our results support and extend previous observations regarding the effects of early life BMI on subsequent MS risk. Observational data supports the concept that BMI during adolescence, particularly late teenage years, is a risk factor for MS^4,7,9,18^. Mendelian randomisation studies have strengthened the notion that this may be a causal relationship^6,7^. It remains debatable whether childhood BMI is a causal risk factor for MS independently of its persistence into adolescence/adulthood, or whether it purely affects MS risk via its strong correlation with BMI in early adult life. In favour of the latter, a recent study used multivariable MR to regress out the effects of genetic instruments for childhood BMI mediated via adult BMI: controlling for the effect of adult BMI abolished the observed MR effect of childhood BMI on MS risk, suggesting that persistently raised BMI into adulthood, rather than transiently increased BMI during childhood, is the causal risk factor^7^. These results, obtained with larger datasets for early life BMI, suggest that what we observe may be driven by the same phenomenon - pleiotropic SNPs acting on MS risk via their effect on BMI in later life rather than in childhood *per se*.

A clear drawback of our study is that we cannot confidently distinguish effects of early life BMI on MS risk that are independent on later-life BMI. Although we employ two approaches to address this question – multivariable MR and exclusion of variants showing nominal association with adult BMI – neither of these approaches is perfect. The high genetic correlation between BMI at different time points, and the relatively small number of genetic instruments (and therefore variance explained in the exposure) raise concerns about the power and the risk of multicollinearity in the multivariable MR estimates. Specifically, the presence of multicollinearity – high correlation between genetic effect estimates of each variant on later childhood BMI and adult BMI – may render the multivariable MR estimates unstable and of low precision. Similarly, our attempts to restrict our analysis to SNPs not associated with adult BMI are limited by the genetic overlap between BMI during early/late childhood and adulthood. As larger GWAS and therefore more genetic instruments become available for childhood BMI at different early life stages, we expect these analyses to become more powerful. Specifically, discovery of variants with strongly ‘time-specific’ effects on BMI are essential to dissect out the causal effect of BMI at different life stages.

A further weakness of our study is the relatively small number of SNPs included in the genetic instruments at each time point. This motivated our approach of collapsing SNPs into time epochs based on shared biology and kinetics of BMI during these time windows, however we recognise that this represents an arbitrary simplification of the data. Although at individual time points our MR instruments are under-powered, our epoch instruments all have good power (F statistics > 10) to detect true effects. The clear null effect of birthweight is confirmed using an external, and better-powered, GWAS^13^. A further possible concern is that population stratification may influence our results, as the exposure GWAS (Norwegian) and outcome GWAS (European) are drawn from slightly different populations. There are three reasons why this is not a major problem for our results. First, the MoBa GWAS were performed on a post-Quality Control subset of individuals who cluster tightly with reference EUR samples from the CEU population (correspondence with authors). Second, these GWAS results show a strong genetic correlation with the ‘comparative body size at age 10’ trait from UK Biobank. Third, allele frequencies for included SNPs are very similar between this GWAS and reference European datasets, such as UKB.

In summary, we have provided evidence from longitudinal Mendelian randomisation that the relationship between early life BMI and MS seems to follow a gradient, with no effect at birth and a gradually increasing influence on later-life MS risk. This gradient may be accounted for by the effect of these genetic variants on BMI in later-life. Phrased differently, we do not find strong evidence that having a higher BMI during childhood which does not persist into adolescence is a causal risk factor for MS. Further work is required to determine whether there is a critical window during which elevated BMI operates as a causal risk factor for MS. Our findings extend previous MR findings in this field and demonstrate the possible value of studying time-varying risk factors with MR.

## Data Availability

We thank MoBa, Prof Stefan Johansson and Marc Vaudel for providing summary statistics for childhood BMI. We thank the IMSGC for providing MS GWAS data. We thank MR-Base for making the TwoSampleMR package available publicly. Download links for datasets used in this study are provided above. All code used in this study is available at https://github.com/benjacobs123456/MR_MOBA_MS.

https://egg-consortium.org

https://imsgc.net/

https://portals.broadinstitute.org/collaboration/giant/index.php/GIANT_consortium_data_files

## Author contributions

BMJ and RD conceived the idea of this work

The primary analysis was performed by LH with support from BMJ

The initial manuscript was drafted by LH with input from BMJ and RD

All authors provided input into critical revisions of the manuscript prior to submission

