## Supplementary Tables and Figures for "Age-specific effects of childhood Body Mass Index on multiple sclerosis risk"

Supplementary table 1: F statistics, Heterogeneity and pleiotropy statistics for epoch-based MR analyses. Exposure refers to BMI during the time window indicated. Mean F statistic refers to the mean of the F statistic calculated for each individual SNP in the instrument. The Egger intercept is the intercept from the MR-Egger regression line. The SE and P value are the standard error of this estimate and the associated p value of the null hypothesis that the intercept=0. Cochran’s Q is the test of heterogeneity for the IVW estimate, DF is the degrees of freedom for the Q statistic, and P is the associated P value for the test of heterogeneity.

| Exposure | Mean F statistic | Egger intercept | SE | Egger P value | Cochran's Q | DF | Q P value |
|---|---|---|---|---|---|---|---|
| Birth – 6 weeks | 56.7 | -0.104404369 | 0.07159 | 0.204545906 | 33.19042805 | 6 | 9.64E-06 |
| 3 months – 1.5 years | 44.0 | -0.006842305 | 0.015081 | 0.656141815 | 14.2349296 | 17 | 0.650408 |
| 2 years – 5 years | 36.9 | -0.103435847 | 0.107708 | 0.438222447 | 0.927124276 | 3 | 0.818878 |
| 7 years – 8 years | 35.3 | -0.085782195 | 0.061427 | 0.29736364 | 3.458673713 | 3 | 0.326162 |

**Supplementary table 2:** results from the leave-one-out analysis. Each line represents the result of the MR analysis for that epoch if the SNP listed in the SNP column were removed. 'All' indicates the overall IVW estimate including all SNPs.

| Exposure | SNP | Beta | SE | P value |
|---|---|---|---|---|
| Birth - 6 weeks | rs11187129 | -0.05557 | 0.132337 | 0.674572 |
| Birth - 6 weeks | rs11708067 | -0.2263 | 0.276029 | 0.412297 |
| Birth - 6 weeks | rs1482853 | -0.2409 | 0.333434 | 0.469993 |
| Birth - 6 weeks | rs2298615 | -0.15031 | 0.273642 | 0.582792 |
| Birth - 6 weeks | rs28642213 | -0.23988 | 0.266935 | 0.368847 |
| Birth - 6 weeks | rs739669 | -0.32243 | 0.26701 | 0.227223 |
| Birth - 6 weeks | rs7772579 | -0.24757 | 0.283141 | 0.381918 |
| Birth - 6 weeks | All | -0.20891 | 0.243413 | 0.390749 |
| 3 months - 1.5 years | rs1032296 | 0.149484 | 0.064047 | 0.019597 |
| 3 months - 1.5 years | rs10493544 | 0.174672 | 0.06423 | 0.006538 |
| 3 months - 1.5 years | rs11676272 | 0.15373 | 0.066713 | 0.021203 |
| 3 months - 1.5 years | rs13038017 | 0.170493 | 0.064192 | 0.007908 |
| 3 months - 1.5 years | rs17145750 | 0.178334 | 0.063764 | 0.005161 |
| 3 months - 1.5 years | rs1772945 | 0.167534 | 0.064396 | 0.009278 |
| 3 months - 1.5 years | rs1820721 | 0.187279 | 0.064651 | 0.00377 |
| 3 months - 1.5 years | rs1985927 | 0.152391 | 0.064085 | 0.01741 |
| 3 months - 1.5 years | rs209421 | 0.167164 | 0.064967 | 0.010081 |
| 3 months - 1.5 years | rs2585058 | 0.16489 | 0.064693 | 0.01081 |
| 3 months - 1.5 years | rs2728641 | 0.150399 | 0.06398 | 0.018737 |
| 3 months - 1.5 years | rs2767486 | 0.138891 | 0.069519 | 0.045729 |
| 3 months - 1.5 years | rs2816985 | 0.145725 | 0.064524 | 0.023917 |
| 3 months - 1.5 years | rs28457693 | 0.154311 | 0.063636 | 0.015312 |
| 3 months - 1.5 years | rs287621 | 0.181764 | 0.064431 | 0.004787 |
| 3 months - 1.5 years | rs3741508 | 0.164378 | 0.064235 | 0.010497 |
| 3 months - 1.5 years | rs6538845 | 0.147865 | 0.064307 | 0.021483 |
| 3 months - 1.5 years | rs6899303 | 0.166645 | 0.064375 | 0.009635 |
| 3 months - 1.5 years | All | 0.162189 | 0.062848 | 0.009861 |
| 2 years - 5 years | rs11676272 | 0.265524 | 0.147335 | 0.071519 |
| 2 years - 5 years | rs12672489 | 0.281904 | 0.139689 | 0.043583 |
| 2 years - 5 years | rs1830890 | 0.310666 | 0.138651 | 0.02505 |
| 2 years - 5 years | rs2767486 | 0.207976 | 0.141361 | 0.141225 |
| 2 years - 5 years | All | 0.267068 | 0.122666 | 0.029467 |
| 7 years - 8 years | rs10493544 | 0.365526 | 0.111862 | 0.001084 |
| 7 years - 8 years | rs11676272 | 0.297798 | 0.15274 | 0.051211 |
| 7 years - 8 years | rs17817288 | 0.255025 | 0.163476 | 0.118757 |
| 7 years - 8 years | rs7132908 | 0.224431 | 0.123289 | 0.068704 |
| 7 years - 8 years | All | 0.289007 | 0.109746 | 0.008453 |

**Supplementary table 3:** SNPs used for multivariable MR analysis. For each epoch, the table shows all SNPs used for the multivariable MR analysis, conditioning on the effect of each variant on adult BMI. The 'epoch' column denotes the age epoch for which the SNP was used as a BMI instrument. 'Beta' refers to the per-allele association with adult BMI ('A-BMI', obtained from the GIANT consortium GWAS), childhood BMI ('C-BMI', from the MOBA GWAS), and MS (from the IMSGC GWAS). P values denote the P value for each of these associations. 'Concordant' refers to whether the SNP beta coefficients for adult and childhood BMI are of the same direction (i.e. both BMI-increasing/decreasing, or with opposite effects on BMI). 'GWAS-significant' refers to whether the SNP achieves genome-wide significance in the MOBA GWAS ('C-BMI') and the GIANT GWAS ('A-BMI'). No SNPs are shown for the 2y-5y epoch as only one SNP was present in both datasets after harmonisation, precluding multivariable MR. For birth – 6w, none of the five SNPs were associated with adult BMI at p<5e-8. For 3mo – 1.5y, two SNPs (*rs10493544* and *rs11676272*) were strongly associated with adult BMI, and had concordant effect directions on both childhood BMI and adult BMI. For the 7y – 8y epoch, all three SNPs were strongly associated with adult BMI, and had concordant effect directions.

| | | Beta | | | P value | | | | GWAS-significant? | |
|---|---|---|---|---|---|---|---|---|---|---|
| Epoch | SNP | A-BMI | C-BMI | MS | A-BMI | C-BMI | MS | concordant | C-BMI | A-BMI |
| Birth – 6w | rs11708067 | -0.0078 | -0.07894 | -1.00E-04 | 5.80E-05 | 5.20E-16 | 0.9973 | Yes | Yes | No |
| Birth – 6w | rs1482853 | 0.0043 | 0.098818 | -0.01511 | 0.016 | 5.90E-32 | 0.3728 | Yes | Yes | No |
| Birth – 6w | rs2298615 | -0.0097 | -0.07128 | 0.045929 | 4.60E-06 | 5.40E-09 | 0.03202 | Yes | Yes | No |
| Birth – 6w | rs739669 | -0.0054 | -0.07157 | -0.02358 | 0.0033 | 4.70E-17 | 0.1841 | Yes | Yes | No |
| Birth – 6w | rs7772579 | -5.00E-04 | 0.065109 | 0.002297 | 0.78 | 5.90E-13 | 0.8997 | No | Yes | No |
| 3 mo – 1.5y | rs1032296 | -0.0069 | 0.052493 | 0.025831 | 9.30E-05 | 1.10E-08 | 0.1316 | No | Yes | No |
| 3 mo – 1.5y | rs10493377 | -0.0078 | 0.056884 | 5.00E-04 | 6.90E-06 | 2.20E-09 | 0.9769 | No | Yes | No |
| 3 mo – 1.5y | rs10493544 | 0.018 | 0.053808 | -0.00638 | 2.80E-28 | 7.10E-09 | 0.6971 | Yes | Yes | Yes |
| 3 mo – 1.5y | rs11676272 | -0.0326 | -0.0887 | -0.0203 | 8.00E-86 | 2.80E-22 | 0.2218 | Yes | Yes | Yes |
| 3 mo – 1.5y | rs13038017 | -0.0036 | -0.05359 | 0.001601 | 0.035 | 1.20E-08 | 0.9229 | Yes | Yes | No |
| 3 mo – 1.5y | rs17145750 | -0.0113 | 0.069787 | -0.02706 | 5.10E-07 | 6.80E-09 | 0.2974 | No | Yes | No |
| 3 mo – 1.5y | rs1772945 | -0.0026 | -0.05626 | -0.0031 | 0.13 | 3.20E-09 | 0.8487 | Yes | Yes | No |
| 3 mo – 1.5y | rs209421 | -9.00E-04 | 0.072738 | 0.006521 | 0.66 | 5.40E-13 | 0.7178 | No | Yes | No |
| 3 mo – 1.5y | rs2268657 | 0.0028 | 0.055523 | 0.023269 | 0.12 | 8.40E-10 | 0.1611 | Yes | Yes | No |
| 3 mo – 1.5y | rs2585058 | 0.0045 | 0.063085 | 0.007373 | 0.0098 | 8.60E-12 | 0.6592 | Yes | Yes | No |
| 3 mo – 1.5y | rs263377 | -0.0051 | -0.05398 | -0.00773 | 0.0041 | 2.90E-08 | 0.6489 | Yes | Yes | No |
| 3 mo – 1.5y | rs2816985 | 0.0012 | -0.05916 | -0.02762 | 0.47 | 5.40E-11 | 0.09259 | No | Yes | No |
| 3 mo – 1.5y | rs287621 | -0.0088 | 0.063684 | -0.0141 | 1.40E-06 | 3.70E-10 | 0.4376 | No | Yes | No |
| 3 mo – 1.5y | rs6899303 | -0.0039 | -0.05715 | -0.00409 | 0.025 | 5.30E-11 | 0.8052 | Yes | Yes | No |
| 7y – 8y | rs10493544 | 0.018 | 0.065159 | -0.00638 | 2.80E-28 | 4.80E-08 | 0.6971 | Yes | Yes | Yes |
| 7y – 8y | rs11676272 | -0.0326 | -0.07817 | -0.0203 | 8.00E-86 | 2.90E-09 | 0.2218 | Yes | Yes | Yes |
| 7y – 8y | rs7132908 | -0.0303 | -0.08135 | -0.03999 | 2.70E-61 | 3.30E-09 | 0.01805 | Yes | Yes | Yes |

**Supplementary table 4:** Multivariable MR results. For each epoch, three MR estimates are shown. The univariable IVW estimate is shown for comparison. Note that the number of SNPs is greater, as some SNPs were not present in the GIANT GWAS summary statistics/were filtered out during harmonisation. 'Multiple' refers to the default multivariable MR method implemented in TwoSampleMR with the *mv_multiple* function. This function fits a weighted multivariable regression model, regressing the associations of each SNP with the outcome on the associations with each exposure. 'Residual' refers to the method describes in Burgess *et al* (see methods).

| Epoch | Method | N SNPs | OR | 95% Lower CI | 95% Upper CI | P value |
|---|---|---|---|---|---|---|
| 3 months to 1.5 years | Inverse variance weighted | 18 | 1.18 | 1.04 | 1.33 | 0.01 |
| 3 months to 1.5 years | Multiple | 14 | 1.11 | 0.97 | 1.26 | 0.13 |
| 3 months to 1.5 years | Residual | 14 | 1.06 | 0.94 | 1.20 | 0.35 |
| 7 years to 8 years | Inverse variance weighted | 4 | 1.34 | 1.08 | 1.66 | 0.01 |
| 7 years to 8 years | Multiple | 3 | 0.45 | 0.05 | 3.92 | 0.47 |
| 7 years to 8 years | Residual | 3 | 0.98 | 0.74 | 1.30 | 0.90 |
| Birth to 6 weeks | Inverse variance weighted | 7 | 0.81 | 0.50 | 1.31 | 0.39 |
| Birth to 6 weeks | Multiple | 5 | 1.09 | 0.66 | 1.81 | 0.74 |
| Birth to 6 weeks | Residual | 5 | 1.03 | 0.80 | 1.34 | 0.80 |

**Supplementary table 5:** MR analyses for each individual BMI time point. Headings are as per table 2.

| Exposure epoch | MR method | Number of SNPs | Beta | SE | P value |
|---|---|---|---|---|---|
| Birth | MR Egger | 6 | 1.187251 | 1.056665999 | 0.324062 |
| Birth | Weighted median | 6 | -0.01469 | 0.147522773 | 0.920676 |
| Birth | Inverse variance weighted | 6 | -0.15031 | 0.273641941 | 0.582792 |
| Birth | Simple mode | 6 | 0.090745 | 0.192853946 | 0.657783 |
| Birth | Weighted mode | 6 | -0.04722 | 0.155153146 | 0.773118 |
| 6w | Wald ratio | 1 | -0.6443 | 0.300492242 | 0.03202 |
| 3m | MR Egger | 8 | 0.397186 | 0.314531414 | 0.253522 |
| 3m | Weighted median | 8 | 0.31016 | 0.119113461 | 0.009217 |
| 3m | Inverse variance weighted | 8 | 0.263156 | 0.093889962 | 0.005066 |
| 3m | Simple mode | 8 | 0.384741 | 0.176421893 | 0.065563 |
| 3m | Weighted mode | 8 | 0.339351 | 0.166334896 | 0.080693 |
| 6m | MR Egger | 11 | 0.316656 | 0.267122985 | 0.266201 |
| 6m | Weighted median | 11 | 0.258437 | 0.106758806 | 0.015488 |
| 6m | Inverse variance weighted | 11 | 0.153684 | 0.078434595 | 0.050067 |
| 6m | Simple mode | 11 | 0.241926 | 0.176372194 | 0.200152 |
| 6m | Weighted mode | 11 | 0.23441 | 0.12384166 | 0.087654 |
| 8m | MR Egger | 11 | 0.444784 | 0.278116586 | 0.144225 |
| 8m | Weighted median | 11 | 0.15731 | 0.102559123 | 0.125068 |
| 8m | Inverse variance weighted | 11 | 0.14013 | 0.077336486 | 0.069994 |
| 8m | Simple mode | 11 | 0.130285 | 0.178961033 | 0.483305 |
| 8m | Weighted mode | 11 | 0.223423 | 0.1411911 | 0.144636 |
| 1.5y | MR Egger | 4 | 0.795002 | 0.445386053 | 0.216192 |
| 1.5y | Weighted median | 4 | 0.284107 | 0.141054541 | 0.043992 |
| 1.5y | Inverse variance weighted | 4 | 0.195237 | 0.126271323 | 0.122063 |
| 1.5y | Simple mode | 4 | 0.279946 | 0.185609437 | 0.228612 |
| 1.5y | Weighted mode | 4 | 0.300814 | 0.173698225 | 0.181732 |
| 1y | MR Egger | 6 | 0.444228 | 0.272698752 | 0.178644 |
| 1y | Weighted median | 6 | 0.242625 | 0.122191032 | 0.047075 |
| 1y | Inverse variance weighted | 6 | 0.199002 | 0.096087102 | 0.038354 |
| 1y | Simple mode | 6 | 0.229002 | 0.170081951 | 0.23598 |
| 1y | Weighted mode | 6 | 0.254779 | 0.128236903 | 0.103676 |
| 2y | MR Egger | 3 | 1.757728 | 2.111127671 | 0.557991 |
| 2y | Weighted median | 3 | 0.27104 | 0.166733736 | 0.104038 |
| 2y | Inverse variance weighted | 3 | 0.310666 | 0.138651211 | 0.02505 |
| 2y | Simple mode | 3 | 0.243996 | 0.180696655 | 0.309423 |

| | | | | | |
|---|---|---|---|---|---|
| 2y | Weighted mode | 3 | 0.248488 | 0.198905785 | 0.337949 |
| 3y | Inverse variance weighted | 2 | 0.212091 | 0.180288002 | 0.239435 |
| 5y | Wald ratio | 1 | 0.281513 | 0.230416856 | 0.2218 |
| 7y | Inverse variance weighted | 2 | 0.213834 | 0.266279589 | 0.42195 |
| 8y | MR Egger | 3 | 0.383833 | 1.270327064 | 0.813196 |
| 8y | Weighted median | 3 | 0.346902 | 0.133083271 | 0.009143 |
| 8y | Inverse variance weighted | 3 | 0.365526 | 0.1118618 | 0.001084 |
| 8y | Simple mode | 3 | 0.322901 | 0.169061037 | 0.196327 |
| 8y | Weighted mode | 3 | 0.33302 | 0.162121183 | 0.176331 |

**Supplementary table 6:** pleiotropy and heterogeneity statistics for individual time point MR. Headings are as for supplementary table 1.

| Exposure | Egger intercept | SE | Egger P value | Cochran's Q | DF | Q P value |
|---|---|---|---|---|---|---|
| Birth | -0.10094 | 0.077359 | 0.26197 | 30.80849 | 5 | 1.02E-05 |
| 3m | -0.00896 | 0.020069 | 0.670911 | 5.591856 | 7 | 0.588128668 |
| 6m | -0.01227 | 0.019174 | 0.538041 | 10.3777 | 10 | 0.408004122 |
| 8m | -0.02179 | 0.019108 | 0.283559 | 8.129033 | 10 | 0.616234734 |
| 1.5y | -0.04823 | 0.034599 | 0.298032 | 3.61715 | 3 | 0.305882942 |
| 1y | -0.01956 | 0.020358 | 0.391025 | 2.470956 | 5 | 0.78086265 |
| 2y | -0.11178 | 0.162726 | 0.616819 | 0.472071 | 2 | 0.789752541 |
| 3y | NA | NA | NA | 0.283546 | 1 | 0.594386108 |
| 7y | NA | NA | NA | 2.65662 | 1 | 0.103119621 |
| 8y | -0.00157 | 0.108607 | 0.99079 | 0.624695 | 2 | 0.731727187 |

**Supplementary figure 1:** scatter plots showing the effect of individual variants used in the MR on standardised BMI (x axis) and MS susceptibility (y axis). The x axis betas reflect the per-allele unit increase in standardised BMI, and the y axis betas reflect the per-allele log odds ratio for MS susceptibility. Error bars reflect 95% confidence intervals. The variants are shown separately for each epoch tested. The fitted lines reflect the IVW estimate, which is a weighted regression line constrained by the intercept.

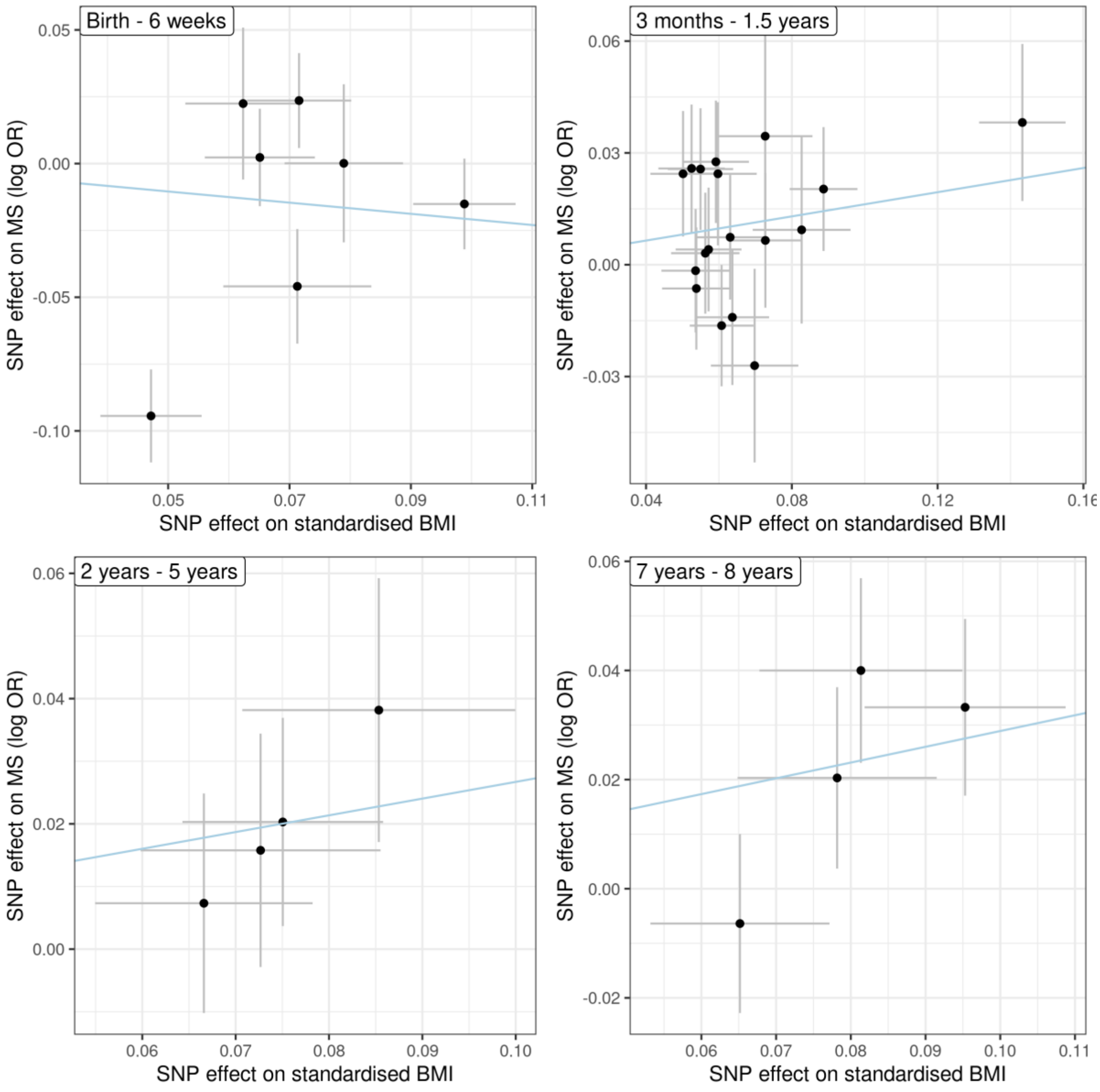

**Supplementary figure 2:** forest plots displaying individual SNP Wald estimates for the effect of BMI on MS risk at each epoch. Individual SNPs used for each instrument are shown on the y axis. The summary measures at the bottom of each plot display the overall MR-Egger and IVW estimates for the effect of BMI during the epoch on MS risk. Points represent point estimates +/- 95% confidence intervals.

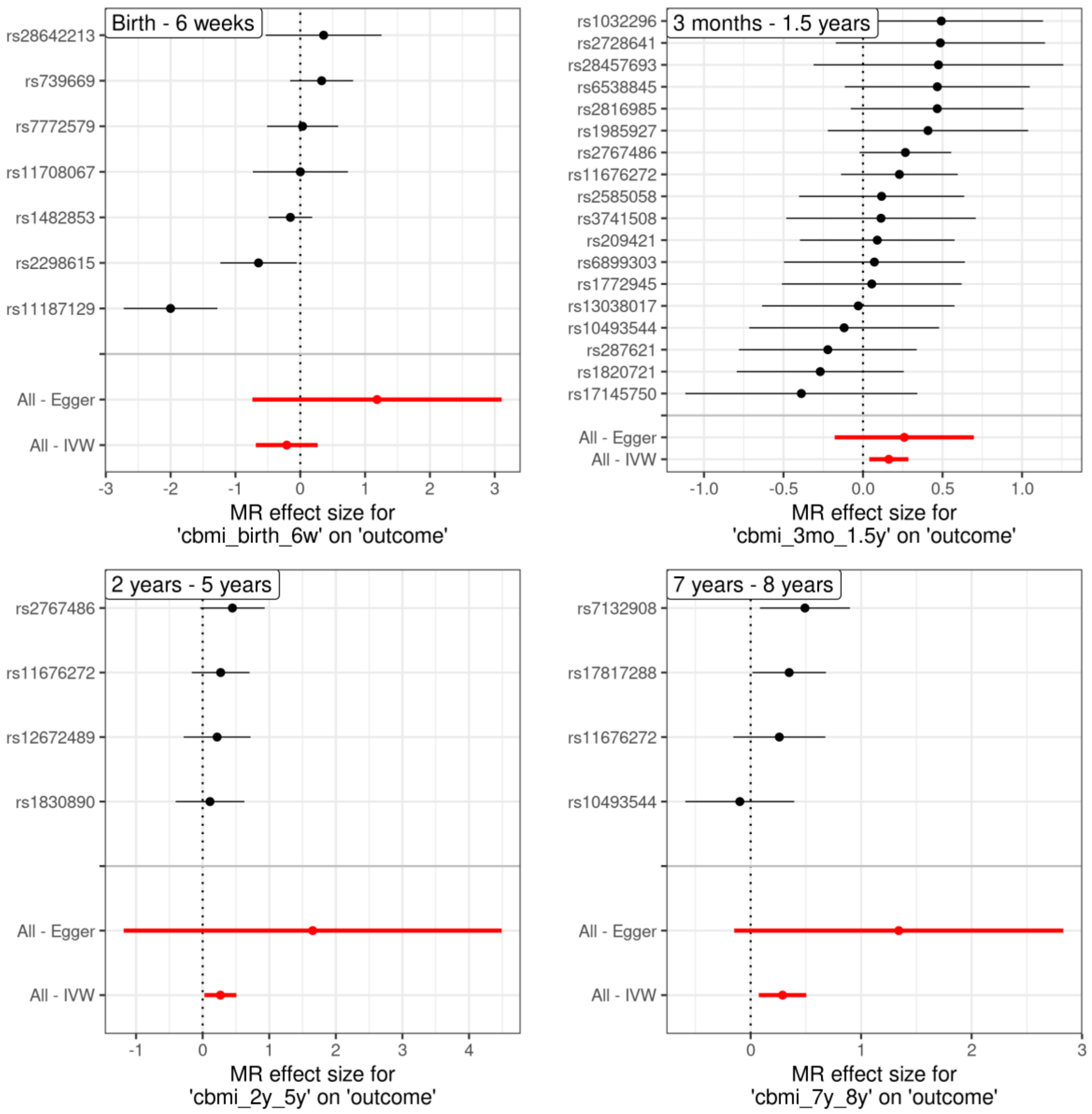

**Supplementary figure 3:** Multivariable MR effect estimates for the effect of BMI at each individual time point on MS risk, accounting for the effect of included variants on adult BMI. For each time point, three effect estimates are shown – "Inverse-Variance Weighted" refers to the univariable MR effect for the time point (i.e. the effect estimates reported in our primary analysis. "Residual" refers to the multivariable MR estimate from a two-step procedure, described in Burgess *et al* (2015), in which associations between SNPs and the secondary exposure (in this case adult BMI) are regressed against the outcome, and the residuals are then regressed against the beta coefficients for the primary exposure of interest, yielding an estimate of the effect of the primary exposure (childhood BMI) on the outcome, conditioning on the effect of the secondary exposure (adult BMI). The y axis shows the epoch of childhood BMI being tested. Individual points represent the MR effect estimates for the effect of a 1 unit increase in standardised BMI on MS susceptibility (on the log odds scale) – multivariable MR estimates represent the effect after conditioning on the instrument's genetic association with adult BMI. At the 2-5y age range insufficient SNPs (n=1) were available to perform multivariable MR.

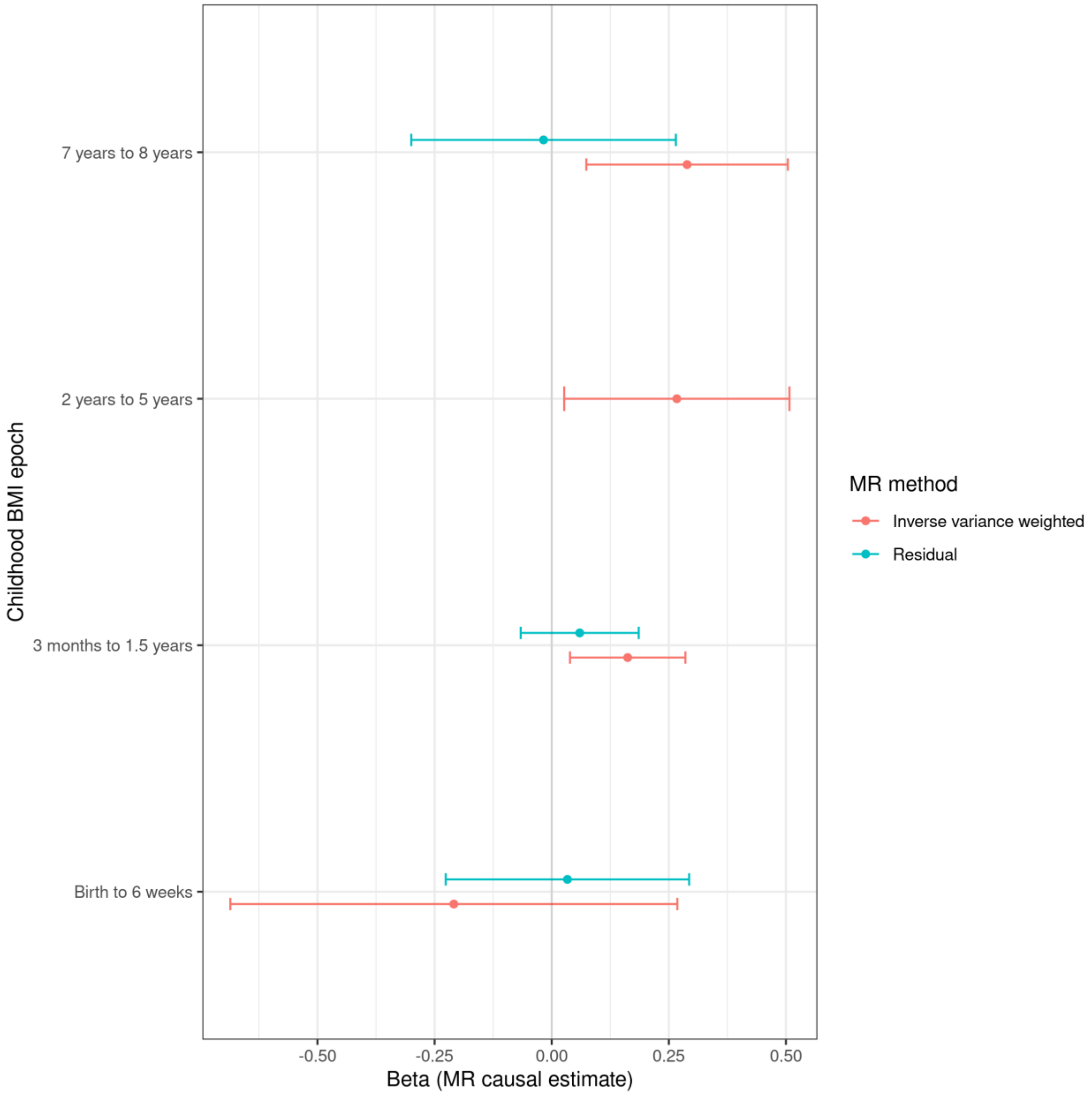

**Supplementary figure 4:** MR effect estimates for the effect of BMI at each individual time point on MS risk. The y axis shows each individual time point for which BMI summary statistics were available. Individual points represent the MR effect estimates for the effect of a 1 unit increase in standardised BMI on MS susceptibility (on the log odds scale). Where enough variants were available, sensitivity analyses were performed and are also displayed. For ages where a single variant was available, only the Wald Ratio is shown.

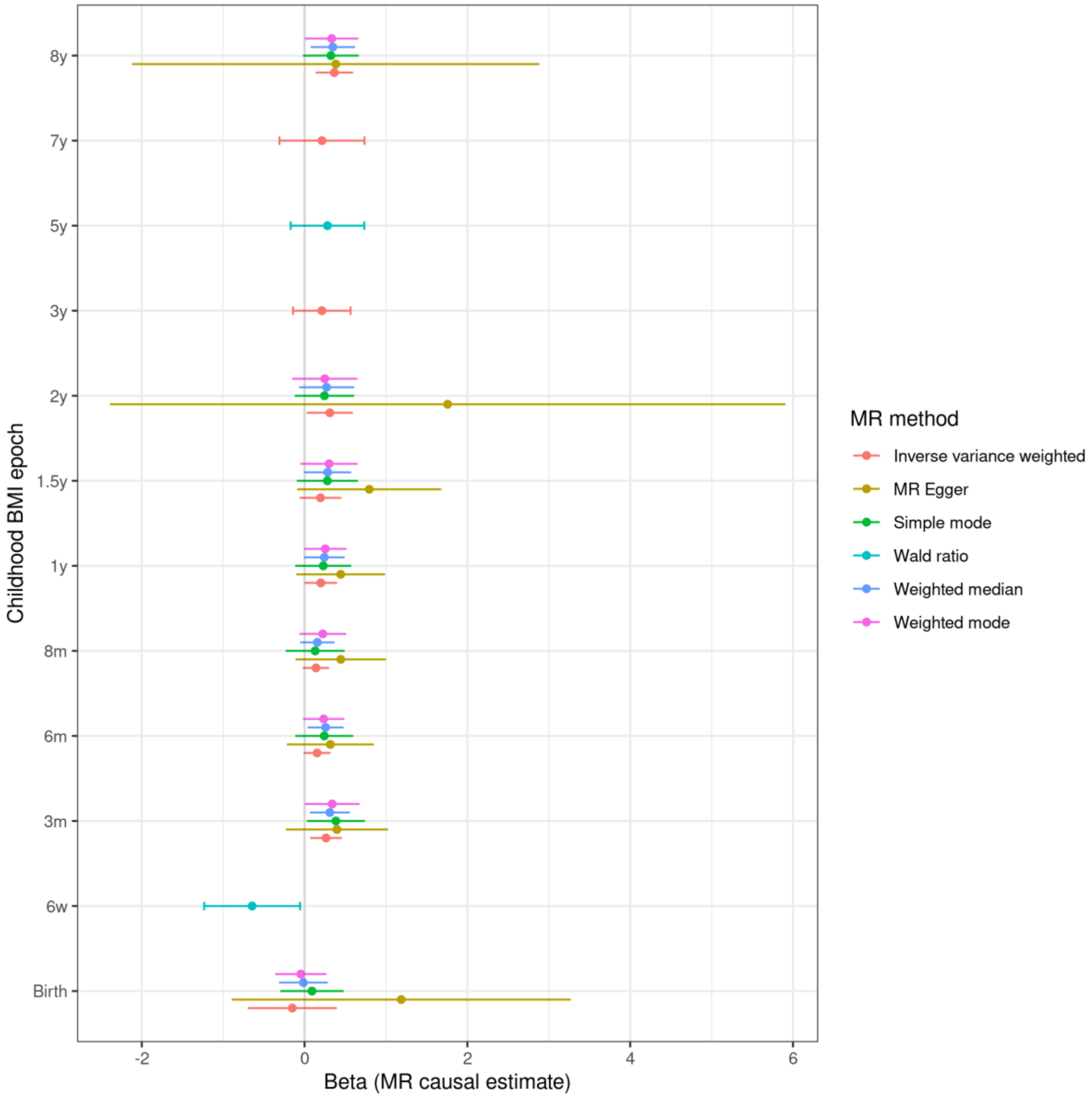
